# Role of Health Equity in the Climate Action Plans of London Boroughs: A Health Policy Report

**DOI:** 10.1101/2023.12.15.23300030

**Authors:** Anandita Pattnaik

## Abstract

**Background:** The World Health Organisation has declared climate change the biggest menace to global health in the 21^st^ century. The health consequences of climate change are well documented. It is also established that vulnerable groups disproportionately bear the effects of climate change. Climate inaction or inequitable climate action can worsen the prevailing health inequalities. Thus, there is an urgent need to implement effective and equitable strategies to minimise the adverse effects and maximise the co-benefits of climate action. The United Kingdom envisions becoming a net-zero carbon country by 2050. The Mayor of London declared a climate emergency in 2018 and aims to make London a carbon-neutral city by 2030. As a result, the London boroughs have published their climate action plans (CAPs) outlining their adaptation and mitigation strategies. But due to a lack of proper guidelines and framework, the plans vary considerably and how health equity is embedded into these documents is currently unknown. This project aims to explore the extent to which health issues are addressed through the CAPs of the London boroughs and if health inequities would be reduced through the greenhouse gas mitigation strategies in the transport sector.

**Methods:** A narrative review of publicly available CAPs of all the London boroughs was conducted to identify if the following 5 health impacts were addressed: food insecurity, vector-borne diseases, respiratory diseases, heat-related, and extreme weather events-related health outcomes. Due to time and resource constraints, health equity implications in vulnerable groups (like the elderly, children, the disabled, and low socioeconomic status) were analysed only in the transport strategies outlined in the CAPs of 10 boroughs. The 3 transport-related strategies – active travel, public transport use, and healthy land use-were selected for major co-benefits. To understand the role of health and equity through climate action, 8 key officials (public health consultants and climate officers) from 7 different local authorities were interviewed. These semi-structured interviews were recorded and thematically analysed using a framework method.

**Findings:** In the 28 CAPs analysed, the health issues were variably addressed. Of the 28 boroughs, 2 mentioned all the health issues listed above, 9 CAPs did not mention any, and the rest noted a few. Most boroughs have focused on equitable transport strategies with maximum health benefits like active travel and other less beneficial options like the electrification of vehicles. But they do not make the best use of communicating the co-benefits. The implications of these transport strategies on vulnerable groups were also variably assessed. The interviews revealed that some councils aimed to improve health and equity through the climate agenda. Still, current practices do not prioritise the role of health in climate action, nor is climate change a public health priority.

**Recommendations:** The recommendations made to the Greater London Authority (GLA) and the local councils are to increase the public health capacity in local climate action, produce climate change related public health evidence, creation of climate change dashboard for public health practitioners, communicate the co-benefits of climate action to the stakeholders, immediate formulation & implementation adaptation strategies, and evaluate the process & impacts of the current CAPs. Further, when developing the CAPs, incorporating ‘Win-Win’ strategies that capitalise on the health and other co-benefits and communicate the economic and wider social gains of the strategies to the public and other stakeholders.

**Limitations:** The main limitation of this report is that only the publicly available CAPs were reviewed; however, there may be the existence of other specific documents (such as air quality or heatwave action plans) which have extensively addressed the health and equity issues. The findings and recommendations are based on the review of the CAPs and interviews conducted. But the evaluation of the implementation of the CAPs was beyond the scope of this report. Further, there is the potential for single researcher bias as the interviews were conducted and analysed by one person.

## **1.** Introduction

### 1.1. Climate change and its impacts on health & equity

As per the World Health Organisation (WHO), equity is the absence of avoidable differences among different groups of people, and health is a fundamental human right. Health equity is achieved when everyone reaches their maximum health and well-being potential.^1^ The WHO has declared climate change as the greatest threat to global health in the 21^st^ century that can directly (through heatwaves, floods, wildfires or storms) or indirectly (through food insecurity or social instability) affect health.^1^ Climate-related health risks disproportionately affect vulnerable groups such as children, women, the elderly, low socio-economic groups, those living with comorbidities, and migrant communities.^1^

As described in the 2010 Marmot Review, climate change poses a catastrophic risk to human health and worsens prevailing health inequities.^2^ Actions directed at mitigating or adapting to the health impacts of climate change would help reduce inequalities. Adaptation is the response to the immediate or expected consequences of climate change. For example, making communities resilient to increasing floods due to sea level rise through structural and non-structural methods. At the same time, mitigation refers to alleviating the longer-term implications of climate change on human health and the natural environment. An example of a climate mitigation strategy to reduce greenhouse gas (GHG) emissions is replacing fossil fuels with cleaner renewable energy sources. Adaptation and mitigation measures are complementary in tackling climate change as the consequences of the climate catastrophe will be felt for decades even after GHG emissions are reduced to zero.^3,4^

Note: Equality is defined as providing the same resources to diverse groups or individuals, while equity means the provision of resources as per the needs of different groups or individuals.^2^ Although there is a significant difference in the meaning between these words, equity/equality and inequity/inequality have been used interchangeably in this report due to inconsistent usage of the word in different documents, and evidence analysed.

### 1.2. Climate action in London, UK

Following the 2015 Paris Agreement of limiting global temperature rise to 1.5°C, the Mayor of London declared a climate emergency in 2018 and has a highly ambitious goal of making London a net-zero carbon city by 2030 (changed from the previous 2050 target).^5^ This is 20 years ahead of the national target of net-zero UK by 2050 and thus requires accelerated action in all domains. Net zero is defined as the anthropogenic carbon generated equals the carbon removed from the atmosphere.^6^ This could be achieved through a combination of measures to reduce GHG emissions and offset the remaining inevitable emissions through activities such as the plantation of trees.

Many London boroughs have thence declared a climate emergency and formulated climate action plans (CAPs). But the CAPs of the London boroughs might vary considerably as there is no formal guiding framework. As mentioned above, most political will around climate action is mitigation related. Adaptation measures, although equally vital, are less emphasised and supported.

### 1.3. Climate action in the transport sector

The road transport sector was the largest emitter of greenhouse gases (31.5% of the total) in the UK in 2021^7^. Due to the COVID-19 pandemic-related restrictions in 2020, GHG emissions from the transport sector had reduced to 30.4% of the total; however, it is rising again as restrictions have eased^7^. The GHG includes carbon dioxide (CO_2_), methane (CH4), nitrous oxide (N2O), and halocarbons.^4^ Besides contributing to GHG emissions, the transport sector is mainly responsible for poor air quality (about 12.4% of PM_2.5_, 33.6% of NO_x_ and other noxious gases), leading to myriads of health impacts.^8^In the UK, 28,000 to 36,000 annual premature deaths are attributable to air pollution.^8^ And London accounted for around 30% (approx. 9400) of these premature deaths, and the associated health services cost between £1.4 and £3.7 billion annually.^9^ Annually, 1 in 6 deaths in the UK is associated with physical inactivity, and the estimated costs are £7.4 billion/annum (with the National Health Services [NHS] accounting for £0.9 billion of that cost).^8^ More than 21,000 traffic related injuries and 96 deaths were reported in London in 2020.^10^ Strategies aimed at reducing the carbon emissions from transport, especially road transport, can also improve local air quality & benefit health while alleviating health inequalities. These have been defined as co-benefits by Intergovernmental Panel on Climate Change (IPCC), i.e., additional positive effects on one objective from the measures or policies directed at achieving another objective.^11^

The mayor’s transport strategy aims that 80% of the journeys in London should be through a combination of active and public transport by 2041.^5^ The objective is to reduce dependency on cars and reduce GHG emissions while improving the health of Londoners. In the last three decades, there have been significant reductions in GHG emissions in sectors such as energy, waste management, and business in the UK. However, the decrement in the transport sector has been minimal, about 2%, from 1990 to 2019 (the GHG emissions peaked in the late 2000s).^7,12^

### 1.4. Rationale for the project

This project seeks to understand the extent to which potential climate-related health impacts are addressed in the CAPs and if health equity is adequately embedded into the transport strategies outlined in the plans. Due to time constrain, only strategies from one sector were considered for health equity consequences. The transport sector was selected due to its enormous potential to reduce GHG emissions, health co-benefits, and minimal improvement over the years (as stated above). Due to a lack of a guiding framework for the local authorities, there is significant variation in the integration of health into the CAPs. Many boroughs have declared a climate emergency while others have not. Even among those who have declared a climate emergency, not all local authorities have a plan to mitigate or adapt to the effects of climate change. Public health is integral to the local authorities’ remit. And it is increasingly important to integrate health into the climate change agenda because of the growing body of evidence and awareness about the impacts of climate change on health. Further, if actions are directed at addressing the health implications of climate change, they would also ameliorate inequalities.^2,13^

A similar study published in 2019 looked at the level of preparedness of local authorities to deal with the health implications of climate change in the Southwest of England^14^, but no such prior project has been conducted in London.

The aims and objectives of this health policy report are outlined in the next section. The methodology is described after that, and the results from the document review and interviews have been summarised. The primary findings of the study and the evidence gathered from other scientific literature have been mentioned, along with the main limitations of the study. Finally, recommendations to the Greater London Authority and the local authorities are enlisted.

## 2. Aims and Objectives^π^

### 2.1 Aim

This report aims to explore the extent to which the climate action plans of the London boroughs address health issues and whether the current strategies aim to reduce health inequities.

### 2.2 Objectives

1. To review the potential health impacts addressed through the CAPs of the different London boroughs.
2. To identify the different transport measures in the CAPs and assess the role of health equity in the key transport measures of a selected sample of London boroughs.
3. To understand the role of health and equity in developing the CAPs by interviewing key local authority officials from a sample of London boroughs.
4. To synthesise the resulting evidence and make recommendations to the Greater London Authority (GLA) and the local authorities to facilitate better incorporation of health and health equity into the CAPs of the local authorities.

π The initial project objectives were to review the CAPs and study the extent to which health impacts have been addressed. This has been modified to present a broader health overview in the CAPs and review the health equity aspects of the strategies in the transport sector.

The title of the health policy report has also been updated to reflect the same.

## 3. Methods

### 3.1 Climate Action Plan Analysis

#### 3.1.1 Search Strategy

A list of all 33 London boroughs was accessed from the London Councils’ website. Then a structured Google search was carried out with the ‘name of the London borough’ plus ‘climate action plan’. Publicly available documents from the websites of the London boroughs were accessed between the 20^th^ to 25^th^ of June 2022.

##### Inclusion criteria

- Boroughs that have a publicly available CAP.

##### Exclusion criteria

- Boroughs that do not that have a publicly available CAP.
- Draft versions of the CAPs uploaded for public feedback, as these might change and are not considered final.
- Boroughs that have different plans that address specific issues such as air quality, heat waves, housing, food etc., but are not integrated into a single climate action document.

### 3.1.2 Data analysis

Using the WHO and UK Government’s climate risk assessments^1,15^, five critical health impacts of climate change that are relevant to the people of London were identified. A narrative review and document analysis of the CAPs was done to determine if the following health impacts had been addressed:

i. Heat-related health outcomes (Physical/Mental trauma)
ii. Extreme weather events (EWE): Flood/drought/wildfire/storm-related health outcomes (Physical/Mental trauma)
iii. Respiratory illness related to air pollution*
iv. Food insecurity
v. Changes in zoonoses/Vector-borne diseases pattern

*There is some overlap between categories (ii) and (iii) above as air pollution can be a consequence of different events such as fossil fuel combustion (transport-related or otherwise), wildfires (becoming more frequent due to climate-related extreme weather), and woodfire burning at homes (often driven by a false belief that it is beneficial for the planet).^13^

Through random sampling, 10 boroughs were selected to review their transport strategies (see Box 1). The sampling was done by numbering each borough from 1 to 28 and then using a random number generator to choose the boroughs. Of all the transport strategies, the 3 most relevant were selected based on the most health benefits gained^16^ and which fall in the realm of the local authorities (See Box 1). The vulnerable groups in the UK were identified for the health equity assessment and are mentioned below (Box 1).^2^

#### Box 1

##### Selected samples of

| Box 1: Selected samples of |  |  |
| --- | --- | --- |
| Boroughs | Transport strategies | Vulnerable groups |
| 1. Brent<br>2. Haringey<br>3. Havering<br>4. Islington<br>5. Lewisham<br>6. Merton<br>7. Newham<br>8. Richmond<br>9. Southwark<br>10. Tower Hamlets | 1. Active travel<br>2. Increased public transport use<br>3. Healthy land use | 1. Older adults<br>2. Children<br>3. People living with a physical or mental disability (Disabled persons)<br>4. Low socioeconomic status |

The data were managed using Microsoft Excel. A narrative review and document analysis were done using a framework informed by health and equity.^17^ The names of the boroughs, health impacts, transport strategies, and vulnerable groups were coded. A total of 3 matrices were constructed to summarise the data in the cells intersecting rows and columns, where each row denoted the code for a borough and the column either denoted the code health impacts or transport strategies or vulnerable groups. This provided an appropriate frame to analyse the different health impacts addressed, the main transport strategies and their effect on vulnerable populations in a deductive manner.

### 3.2 Semi-Structured Interviews

#### 3.2.1 Recruitment of participants

Participants were recruited with support from the GLA. A convenience sample of officials working in the local authorities, consisting of public health consultants and climate officers, were invited to participate in the study via direct personalised emails or bulk emails through the GLA network. A total of 6 public health consultants (PHCs) and 2 climate officers (Climate officers [COs] is a generic term being used here to refer to exclusive job profiles within the local authorities that manage climate action projects) from 7 different London boroughs responded to the emails. They were provided further details in the form of a participant information sheet and a consent form. The names of the boroughs represented through the interviews are not included in this report to ensure that the anonymity and confidentiality of the participants are maintained. However, the sample consists of a mix of inner and outer boroughs from the north, north-west, south-west, south, central, and east of London.

#### 3.2.2 Conducting interviews

Interviews were conducted between the 26^th^ of July 2022 and the 5^th^ of August 2022. A topic guide was developed based on the objectives with appropriate cues to prompt more information during the semi-structured interviews. Individual interviews (except for participants 5 & 6, who represented the same borough and wanted to be interviewed together) were conducted virtually and recorded by the researcher. The recorded videos were transcribed manually. Field notes reflecting the general rapport, questions asked by the participants, and emerging topics were made after each interview.

#### 3.2.3 Analysis of the qualitative data

All the data were managed using Microsoft Excel and Microsoft Word.

The participants were randomly coded from P1 to P7 (Except for P5 and P6, who were jointly interviewed). Ritchie and Spencer’s framework method was used to analyse the qualitative data.^18^ Through a deductive approach, a set of initial key themes were developed and coded based on the objectives/topic guide. Additional themes and sub-themes were identified and coded inductively as they emerged from the data. The list of themes and sub themes codes were applied to the data. A table was created to categorise the codes and the data. The table was analysed for patterns across and within data.

### 3.3 Ethics

This research study was approved by the London School of Hygiene and Tropical Medicine MSc Ethics Committee (Ref: 27244)

## 4. Results

### 4.1 Analysis of health impacts in the climate action plans

Out of the 33 London boroughs, 28 boroughs had a publicly available CAP and were included for analysis. The boroughs of Barnet, Bexley, Bromley, and Hackney, were excluded as they did not have a publicly available CAP, while Greenwich only had a draft version of their CAP. As presented in Table 1, many boroughs outlined the different health risks due to climate change in the CAPs. Only 2 out of the 28 boroughs mentioned all the 5 health risks, while 9 boroughs did not mention any of the health risks listed above. However, none of the boroughs has adequately quantified or holistically described these health issues.

**Table 1:** Imminent health risks addressed by the CAPs of 28 London boroughs.

| London Boroughs | B & D<sup>26</sup> | Brent<sup>22</sup> | Camden<sup>27</sup> | City of London<sup>25</sup> | Westminster<sup>28</sup> | Croydon<sup>29,30</sup> | Ealing<sup>24</sup> | Enfield<sup>31</sup> | H & F<sup>32</sup> | Haringey<sup>33</sup> | Harrow<sup>34</sup> | Havering<sup>35</sup> | Hillingdon<sup>19</sup> | Hounslow<sup>36</sup> | Islington<sup>6</sup> | K & C<sup>37</sup> | Kingston<sup>38</sup> | Lambeth<sup>20</sup> | Lewisham<sup>39</sup> | Merton<sup>40</sup> | Newham<sup>41</sup> | Redbridge<sup>42</sup> | Richmond<sup>21</sup> | Southwark<sup>43</sup> | Sutton<sup>44</sup> | Tower Hamlets<sup>45</sup> | Waltham Forest<sup>46</sup> | Wandsworth<sup>47</sup> |
| --- | --- | --- | --- | --- | --- | --- | --- | --- | --- | --- | --- | --- | --- | --- | --- | --- | --- | --- | --- | --- | --- | --- | --- | --- | --- | --- | --- | --- |
| Heat-related health outcomes |  | X |  | X | X |  |  | X | X |  |  | X | X |  |  |  |  | X |  |  |  | X | X | X |  |  | X |  |
| Respiratory illness* | X |  |  |  |  |  |  |  | X |  |  | X | X |  |  |  |  | X |  |  |  |  | X | X | X | X |  |  |
| EWE: F/D/W/S related health outcomes |  | X |  |  |  |  |  |  | X |  |  | X |  |  |  |  |  |  |  |  |  |  | X |  |  |  |  |  |
| Food insecurity | X | X | X | X |  |  | X |  | X |  |  | X | X |  |  | X |  | X |  |  |  |  | X | X |  |  |  |  |
| Vector-borne diseases |  |  |  | X | X |  |  |  | X |  |  |  | X |  |  |  |  | X | X |  |  |  | X | X |  |  |  |  |

\* Barking & Dagenham
\*\* Hammersmith & Fulham
\*\*\* Kensington & Chelsea

The majority of boroughs have highlighted the health impacts of rising temperatures and more frequent heatwaves, mainly in terms of increased mortality. Boroughs of Hillingdon, Lambeth, and Richmond have elaborated that vulnerable populations such as the elderly, infants, and those with underlying illnesses are disproportionately affected by such extreme heat events.^19–21^ Hillingdon council have additionally considered increased violent behaviour and aggression due to high environmental temperatures adversely impacting mental health^19^. Most boroughs have the creation of greenspaces on their agenda, which would provide cool spots and increase resilience to such events.

The borough of Richmond has highlighted the short- and long-term implications of exposure to poor air quality.^21^ This is vital as the long-term health impacts are often not tangible, making it difficult to comprehend. Lambeth has acknowledged the broader economic benefits of health improvements through air pollution mitigation strategies. For example, increased productivity due to reduced school and work absenteeism and decreased healthcare costs due to a decrease in the incidence of asthma or cardiovascular disease.^20^

EWEs such as flood/drought/storm/wildfire will become more severe and frequent as climate change progresses. Only 4 councils, however, have mentioned how such events would adversely impact health. The borough of Brent has written about the mental health impacts such as depression, anxiety, and post-traumatic stress disorders associated with such events.^22^ The CAP of Richmond states that the contamination of surface water with sewage leads to infectious disease outbreaks.^21^ Only the borough of Lambeth has mentioned climate anxiety^20^, which is the feeling of distress due to concerns about the effects of climate change. There is growing evidence of climate anxiety in children and young adults, primarily due to perceptions of government inaction in responding to climate change.^23^ Hence, local councils must have a CAP that adequately communicates the various climate mitigation and adaptation strategies to the community.

Food insecurity is a consequence of climate change-related changing weather patterns, water shortage, and crop failures leading to global supply chain breakdown. The borough of Ealing elaborates that this results in rising food prices, causing food poverty that manifests as malnutrition in all its forms.^24^ Most boroughs have mentioned communal food cultivation to reduce carbon emissions and food wastage, but if done at a large scale, it can act as a buffer when there is a surge in food prices. As climate change progresses and there is a further rise in temperature and changing weather patterns, vector-borne diseases and zoonoses will emerge. This has been identified as a public health problem by 8 boroughs, but only the City of London has a strategy to prevent outbreaks by surveillance through horizontal scanning of infectious diseases and early warning systems.^25^

### 4.2 Analysis of health equity in the transport strategies

As mentioned above, the following transport strategies with the greatest health co-benefits due to improvement in air quality and increased physical activity were selected for review:

1. Active travel
2. Increased public transport use
3. Healthy land usage

Of the 10 boroughs randomly selected, 9 boroughs have mentioned active travel combined with public transport use as the primary mode of transportation. The actions aim to significantly increase active travel, i.e., walking and cycling in the boroughs. Innovative bike hiring schemes and providing lessons on riding bicycles to encourage residents to take up cycling have been stated. Some boroughs have planned the creation of cycling clubs and trips to champion this sustainable behaviour. Additionally, the reallocation of road spaces to increase cycling infrastructure and pedestrian areas to create an environment that induces active travel in the neighbourhood is planned.

Expansion of public transport networks, especially buses, to increase accessibility throughout the boroughs while also transiting to renewable sources of fuel is necessary. This can be achieved by lobbying and collaborating with Transport for London (TfL). The availability of rapid, affordable, and accessible public transport can prompt people to use these services instead of private vehicles. Especially when combined with private car ownership disincentivising schemes such as parking charge levy, registration taxes, and the creation of low-traffic neighbourhoods. Creating low-traffic areas fosters public transport and active travel as a natural mode of transportation. Out of the 10 boroughs, 7 have mentioned developing such neighbourhoods that are either car-free or lite.

Additionally, most of the boroughs have mentioned electrification of the fleet and expansion of the infrastructure to support electric vehicles. However, the borough of Tower Hamlets has focused on the electrification of vehicles as the primary transport strategy without much emphasis on active travel or public transport utilisation.^45^ This could lead to worsening inequalities, as the high cost of electric vehicles makes it difficult to afford for poorer households.

The documents were reviewed to identify whether the councils considered the effect of the transport strategies (as mentioned earlier) on different vulnerable groups such as the elderly, children, disabled persons, and low socioeconomic status. Although 8 out of 10 boroughs have mentioned that climate change disproportionally affects vulnerable populations, only 5 of the boroughs have specified the implications of any transport strategy on these groups (See Table 2).

**Table 2:** The impact of the transport strategies on different vulnerable groups assessed by the local councils.

| London boroughs | Brent* | Haringey | Havering | Islington | Lewisham | Merton | Newham | Richmond | Southwark | Tower Hamlets |
| --- | --- | --- | --- | --- | --- | --- | --- | --- | --- | --- |
| Elderly | X |  |  |  |  |  |  |  |  |  |
| Children | X |  |  |  |  |  |  | X |  |  |
| Living with a disability | X |  |  | X | X |  |  | X |  |  |
| Low SE groups |  |  |  |  | X |  |  |  |  |  |

\*Brent had a separate equality impact assessment document available right below the CAP on the same webpage.

The borough of Brent has mentioned an increased risk of dementia in the elderly and the development or exacerbation of asthma in children due to high levels of air pollution. Further, chronic childhood exposure to the pollutants in the air can lead to lung cancer in adulthood. Active travel can help reduce air pollution levels and thereby ameliorate the health risks for the elderly and children. Additionally, transport strategies to reduce air pollution near schools would help to reduce the adverse health effects experienced by children.^22^ While Richmond council has explored the potential to reduce childhood obesity through active travel by encouraging the use of cycles, skating, and scooters.^21^

The boroughs of Brent, Islington, Lewisham, and Richmond have considered the impact of active travel and public transport use on people living with disabilities.^6,21,22,39^ For example, actions to incentivise active travel may have a negative impact on individuals with disabilities, who may be less able to travel by walking or cycling. Also, not all public transport options are fully accessible for disabled individuals. Additional special considerations include flat road crossing, clear pavements, and increased resting spots along popular routes such as those connecting stations and town centres were stated.

For low socio-economic groups, expanding the public transport network to increase accessibility would be advantageous, as has been mentioned by the borough of Lewisham.^39^

### 4.3 Semi-structured interviews

A total of 8 participants (6 PHCs and 2 COs) from 7 different London boroughs were interviewed.

#### 4.3.1 Role of health in the CAPs

Most participants said that there is a definite role of health in the CAPs and that the local councils aim to improve health and wellbeing through climate action. But a majority of the interviewees agreed that the health narrative is not well reflected in the CAPs. A few participants mentioned that climate change is also not a public health priority.

> “For many people, it’s not immediately intuitive that health and public health has something to add when we talk about climate change. There is not even an attempt to make the best use of the health argument within the context of climate action. They talk about sustainability and equality in terms of providing the less wealthy who do not have private gardens access to communal green spaces. But they don’t go one step further to talk about the health co-benefits.” (P1)

> “Climate specialists will focus on net-zero; that makes sense. But there is an opportunity to have health core benefits. Climate people measure their worth from just net zero, but that means missing out on many other benefits. If we get so many people active, we can model, and we then prevent so many people from becoming obese. We then prevent so many people from getting diabetes. And we further prevent many people from getting an amputation, going blind, or needing a kidney transplant. We are not very good at doing that across different sectors. So, the net zero people model their net zero. They don’t link all this up. Therefore, investment hardly ever goes across and cost-benefit analysis that goes across.” (P3)

#### 4.3.2 Barriers to implementing actions built on health equity

Although the boroughs thrive to ensure social justice and reduce health inequalities, all the participants interviewed stated budget cuts for the local authorities to be a hindrance to equitable climate action with health at its core.

> “Funding is a big barrier because the scale of what we are trying to do is enormous. Our budget is reducing, and it would reduce even further in the next few years.” (P5)

> “Struggling to resource existing priorities like homelessness, housing, older residents etc. Our budgets have reduced significantly over the years. Climate emergency is a massive expectation but has no associated funding.” (P7)

A few participants mentioned innovative funding methods, such as green schemes. Interestingly, one of the participants pointed out that the concept of ‘matched funding’, in a way, perpetuates inequalities. Matched funding is a financial mechanism in which an organisation pledges to pay a sum of money equivalent to that generated through other means.^48^

> “What you find is that people who have lots of money match funds for what they like. So, that’s one of the approaches that does tend to perpetuate inequalities.” (P6)

Most participants identified a lack of capacity and competing priorities as another significant barrier. Often, climate action takes a back seat as officials deal with other concerns such as cost-of-living crises or the COVID-19 pandemic.

> “Time and capacity are a barrier. Competing priorities, which are more acute and need attention right now. But it’s getting better; now, there’s a team dedicated to climate action. It’s a part of my role, although a tiny part. In the future, we may have a dedicated person in every team that contributes to climate action. For example, if it were 50% of my role instead of 1/4th of my role, I’d be able to do more on that.” (P4)

> “Climate change is not an immediate priority; it’s sort of negotiable. Once we have got the cost-of-living crises done, then we do climate change. First, there was COVID; while we tackled COVID, we couldn’t think of climate change.” (P3)

Engaging, influencing, and inspiring the community to get on board with climate action is another challenge commonly faced by local authorities. On a superficial level, there is community support, but when measures are implemented, it becomes inconvenient for people leading to dissatisfaction and opposition.

> “We’ve got many affluent people who own big four-by-four vehicles, and they don’t want to get out of their vehicles and walk or cycle. So how do we do that? How do we make things really easy so that it’s convenient to make the right choices? And it is not just about telling people what to do. It has to be easy. It has to be convenient.” (P8)

> “I’ve talked about public buy-in. I think that’s a real barrier because sometimes things seem okay on the surface when you’re talking at quite a high level, but once you start to get into the details of what changes are needed, that’s where people don’t like it anymore. So, a classic example of this is parking. Many people might say that they’re worried about the environment or want to live in a more healthy and pleasant neighbourhood. But one of the answers to that is to reduce car use and to make streets quieter. And then the minute you try to take away parking or charge for parking or things that will disincentivise driving, people object.” (P2)

Most boroughs conduct an equality impact assessment to evaluate the potential consequences of the policy actions on vulnerable groups. However, the process is quite tedious and cumbersome; hence, it is more often a formality rather than an endeavour to identify and mitigate any resulting adversities.

> “If you’re doing a proper equality impact assessment, it takes a lot of time, and these tools then become sort of a tick box rather than anything else. It comes down to capacity and priority. So, if equality impact assessment is implemented in letter and spirit for climate change agenda and ensuring the health impact is carefully considered for any strategy or policy, it should be done properly with consultation with all those sectors. Some of the policies are rushed because of pressure from politicians or pressure from different stakeholders. So then, although these tools exist, the equality impact assessment, and others, they’re not overlooked, but they are also hurried through, just to tick boxes.” (P4)

#### 4.3.3 Scope for improvement

The participants mentioned that climate change should be made a public health priority. There is an urgent need for a guiding framework and precise set of guidelines that could ensure appropriate integration of health into climate action so that the strategies are equitable. For example, climate action should be included in the Public Health Outcomes Framework along with upscaling the knowledge about the health impacts of climate change amongst public health officials.

> “I think that knowledge and skills gap is a real barrier. Many people don’t, either don’t quite know what the issues are, or they don’t feel confident to address them. It’s always helpful to have the right knowledge and tools. And because the climate is relatively new to lots of people and climate impacts and how you can tackle climate change. That’s a real gap for lots of people. So having some guidance, tools, frameworks, and things like that are useful.” (P2)

> “Climate action is not really a priority for public health. So, the public health outcome framework of the UK does not include climate action. It does include air quality. This is something we, as a system, need to lobby for so that the directors of public health have an explicit role in climate action.” (P1)

Most interviewees agreed that there is an urgent need for robust adaptation plans at the local councils’ level as there is a large gap between mitigation and adaptation action.

> “I think we need adaptation urgently now. It’s a bit like in health service, you do treatment, and you do prevention. So, the treatment is almost like the adaptation; it’s something now. And then mitigation is a bit like prevention. You can’t just say…ʽOh! Sorry, you die of a heart attack, but the next one gets smoking cessation advice.’. No, you have to do both.” (P3)

Half of the participants mentioned that developing a joint strategic needs assessment (JSNA) for climate change would help put health equity at the core of climate action. The JSNA is a systematic needs assessment process by the local authorities to identify the local communities’ current and future health and healthcare service requirements to inform decision-making. JSNA for climate change has already been done by the boroughs of Brent, Lambeth, and Southwark. But it is not the norm.

> “…(We need to) have an explicit mention of climate change within the JSNA.” (P1)

> “(Speaking about JSNAs) It’s that evidence-based stuff. I think that in the public health world, there’s been quite a lot that’s gone into producing evidence-based documents around our health priorities. Some of it has already happened around air quality, so it probably isn’t that difficult to do. We need that for climate change.” (P7)

## 5. Discussion and Recommendations

### 5.1 Key findings

The extent to which climate-related health issues are addressed in the CAPs of different London boroughs varies. Most of the boroughs focused on equitable transport strategies, but there is scope to become more comprehensive in terms of achieving the most significant health, equity, and economic co-benefits. The interviews with key officials from local authorities also revealed that the boroughs aim to improve health and reduce inequalities, but currently, they do not form the backbone of climate action. Public health needs to be prioritised in climate action and vice versa; climate change needs to be a public health priority.

### 5.2 Health & climate action

The document reviews and the qualitative data analysis revealed a need for WHO ‘Health in all policies’ approach in the CAPs. Actions directed to mitigate or adapt to climate change have a myriad of co-benefits.^11,16,49^ Climate change is such a complex and dynamic process that the efforts to tackle it need to be integrated across different sectors. This would help evaluate the co-benefits associated with various climate actions, and investments could be utilised to prioritise strategies with maximum health and other co-benefits. Interestingly the borough of Lambeth mentions climate anxiety which highlights the fundamental need for the local councils to develop CAPs and communicate the different strategies with different stakeholders.^20,23^ Additionally, the community should be regularly notified about the progress in the implementation of the CAPs. This would help create an optimistic aura around climate talk, which is mostly about gloom and doom at the moment, as mentioned by some interviewees.

The Mayor of London aims to make the city net zero by 2030; however, many boroughs aim to be net zero by 2041 and a few by 2050. There is a need for alignment of action at the local, regional, and national levels to be a net-zero economy. One of the reasons for the misalignment could be that the London net-zero target was recently changed from 2050 to 2030. To achieve this extremely ambitious goal, the national & regional authorities need to support local action by providing sufficient funds, tools & resources and building capacity. All participants stated that dwindling local authority funding was a major barrier to incorporating health in the CAPs. This is consistent with the findings of the Marmot Review follow-up report published in 2020 that there has been a significant reduction in funding to local authorities^2^. Additionally, the implementation of the CAPs has been severely disrupted due to the COVID-19 pandemic.^22^

Climate change is currently not a public health priority in the UK, as mentioned by many of the participants in the study. For example, the Public Health Outcomes Framework (PHOF) does not include climate change. PHOF is an overarching vision for public health with a set of outcomes and indicators that help determine how public health is improving in a region. It is reviewed and updated every 3 years, the last update being in 2019.^50^ Many interviewees stated that although there are increased discussions about climate amongst the directorate of public health, the inclusion of climate change-related health outcomes/indicators into the PHOF will ensure appropriate integration of health equity into the climate action agenda. This is because the PHOF aims to reduce health inequalities, and there is strong evidence that climate change would exacerbate existing health inequalities.^2^ Additionally, the Joint Strategic Needs Assessments (JSNAs), commissioned by the local authorities to provide evidence for action to improve health and wellbeing within the local communities^51^, should be conducted for climate change. Three boroughs in London have already done this, but it would be systematised if all the local councils produced a JSNA, as it would further strengthen the role of health equity in climate action strategies.

Currently, action directed through CAPs is mostly net-zero-related mitigation strategies. The adaptation strategies are inadequate and fragmented. There is a dire need to augment the adaptation strategies in London from a public health viewpoint. A tool developed by the University of Exeter called Local Climate Adaptation Tool (LCAT) provides evidence-based adaptation recommendations to deal with the different effects of climate change and improve the health of the population.^52^ However, it is only at a prototype stage and is available only for the Cornwall region in the UK. Expansion of such a tool can conspicuously improve climate adaptation action from a public health perspective. A need for the expedition of adaptation action was also felt during the heatwaves in the summer of 2022 when the UK experienced record-breaking temperatures of over 40°C for the first time in climate history.^53^ This alarming consequence of global warming has created an ‘Overton Window’ for formulating and implementing climate policies that are acceptable to the mainstream population.^54^ It is also an opportune moment to engage different stakeholders in the climate action agenda and encourage the inculcation of sustainable behaviours as they perceive the immediate threat of climate change.^55^ Heat-related morbidity and mortality are directly proportional to a rise in global temperature, and with frequent exposure to extreme heat, the burden of cardiovascular (e.g., myocardial infarction), respiratory (e.g., infectious diseases and exacerbation of asthma), and neurological (e.g., heat stroke related) sequelae increases.^56,57^ There are also concerns about a surge in the cases of skin burns and cancer from increased exposure to UV light with rising temperatures.^4,57^

Extreme weather events (EWEs) such as floods, droughts, cyclones, and wildfires will become more frequent due to global warming, rising levels of seawater, and changes in precipitation patterns. Such events have a direct (e.g., drowning due to flooding) and indirect (e.g., malnutrition or exposure to environmental toxins) adverse impact on human health. An increment in water- and food-borne diseases such as diarrhoea due to Escherichia coli (E.Coli) is expected with a rise in EWEs, especially due to flash floods and cyclones.^57^ These events are detrimental to mental health, and the outcomes manifest in various forms, such as post-traumatic stress disorder, depression, anxiety, and psychological distress. There is also a proportionate association between alcohol consumption and psychotropic drug use with EWEs. The burden on health care systems is significant with frequent extreme climatic events.^56,57^ Further, increased precipitation and temperature can result in the rise of vector-borne diseases in temperate countries like the UK. Mosquito-borne infections such as the West-Nile virus, dengue, chikungunya, and malaria will become more prevalent in the future.^4,56,58^ There is also a possibility of an increase in tick-borne diseases like Lyme disease and tick-borne encephalitis.^57,58^ It is worthwhile to note that while the expansion of greenspaces has a multitude of health co-benefits, there is a risk of an increase in vector borne diseases due to the formation of breeding sites for these organisms.^59^ Hence, greenspaces must be meticulously designed to prevent the creation of thriving environments for these vectors. EWEs are also associated with crop failures and food shortages. Food security is defined by the United Nations (UN) as access to affordable, safe, and nutritious food in sufficient quantities for every individual at all times.^60^ As food is a global commodity, if its production is affected due to different climate change-related events in one part of the world, it can affect food consumption in another part of the globe. Food shortages can lead to rising prices, adversely influencing consumer behaviour towards unhealthy food choices. This can further exacerbate health inequalities by worsening nutritional deficiencies and increasing the prevalence of obesity and related diseases, especially in children, young adults, the elderly, low-socioeconomic and other disadvantaged groups.^60^ This is primarily due to easy accessibility to inexpensive, nutrient-poor, and obesogenic foods. The CAPs of most London boroughs do not sufficiently address the mortality and morbidities associated with the complex and interlinked climate-related events.

The CAPs of the London boroughs are quite aspirational, and to achieve their climate goals, these plans must be aggressively implemented. And for this, there is an urgent need to escalate funding, increase capacity and upskill the public health task force in climate action. Both process and impact evaluation need to be done to assess if the objectives of CAPs have been achieved.

### 5.3 Health equity in the transport strategies

In 2019, 25.75% of the global CO2 emissions could be attributed to the transport sector.^12^ Following year with the COVID-19 pandemic, there was a reported decrease in GHG emissions from the transport sector. However, it is rising again as the world is reverting to the old ways of living.^7^ There is potential for colossal emissions reduction from this sector, but efforts to achieve this have been meagre despite strong evidence for action. Equitable transport strategies can directly improve population health and positively influence the social determinants of health in terms of better access to employment, education, health care, community services and others.^11,16,49^

Transport strategies to mitigate GHG emissions that have health co-benefits:

1. Active travel – Walking or cycling has almost zero emissions and maximum health benefits through increased physical activity and improved air quality.^4,16,49,61^ Increased physical activity can reduce obesity and prevent type 2 diabetes, heart diseases, stroke, hypertension, breast cancer, colorectal cancer, and other obesity related co-morbidities.^4,16,62^ Studies have shown that a median increase of active transport from 4 mins to 22 mins can reduce the risk of diabetes and cardiovascular diseases by 14%.^4^ Physical activity also helps to improve mental health and cognitive functions.^16^ Evidence suggests that within 20 years, there will be savings worth approximately £17 billion for the NHS due to a reduced prevalence of diseases associated with physical inactivity.^62^ A modal shift to active travel is associated with improved air quality. The decrease in fossil fuel use leads to a fall in noxious emissions from motorised vehicles, such as nitrogen oxides (NO_x_), carbon monoxide, and particulate matter (PM_2.5_ and PM_10_).^16,49^ Of all the air pollutants, the most harm to human health is from ambient particulate matter, particularly PM_2.5_, which can penetrate deep into the lungs to reach the alveoli and the bloodstream. Even within a city, regional and local variations in the PM_2.5_ levels can differentially impact populations & their health. Air pollution has a myriad of detrimental health effects. It affects the cardiovascular system (such as the increased risk of heart attacks and heart failure), respiratory system (such as exacerbation of chronic obstructive pulmonary disease [COPD] & asthma, increased risk of lung cancer and respiratory tract infections), neurological system (such as the increased risk of stroke and Alzheimer’s disease) and other conditions (such as autism spectrum disorders, premature birth, and low birth weight).^16^ Yet, air pollution due to other sources, such as woodfire burning at homes or more frequent wildfires due to climate change, would continue to be a health hazard. In addition, a significant transition to active travel results in a diminished number of motorised vehicles on roads which is associated with a reduction in noise pollution. This alleviates noise-related morbidity (chronic annoyance, sleep disturbance, and ischemic heart diseases) and mortality (12,000 premature deaths/year in Europe).^63^
2. Rapid, accessible, and affordable public transport network – Buses and trains powered by petrol or diesel have a lower GHG emission per person than conventional fuel-powered cars and similar/less than electric cars^16^. Environmentally friendly public transport that runs on renewable energy sources has substantially lower emissions than private electric vehicles. Increased public transport use could improve the air quality due to diminished personal vehicle use.^16^ Also, mild-moderate physical activity (on average 21 minutes/day) is involved when using public transport. This benefits health as the recommended level of weekly exercise is inadvertently achieved through the utilisation of public transportation.^64^
3. Smart redesigning of urban form – Decrease in urban sprawl and creation of compact neighbourhoods (such as 15-minute neighbourhoods) in which essential amenities such as supermarkets, schools, or places of work are within walking distance.^16,65,66^ Along with an intricate public transport network, this would minimise the trade-offs that would otherwise arise with the reduction in private vehicle use. Such urban designs with abundant green spaces will help promote active travel and improve life expectancy and mental health outcomes.^16^ Green spaces promote greater resilience to climate change-related events such as global warming, heat waves, and urban heat islands through natural cooling mechanisms.^16^ One such example is the ‘Superblocks’ in Barcelona which has people-centred urban planning. It has redefined pedestrian mobility by prohibiting motorised vehicles from entering the inner part of the blocks, with certain exceptions like ambulances or goods delivery vehicles. It creates an optimal environment for active travel to thrive.^67^ It is a scalable and sustainable intervention that could be expanded to other megacities. Traffic related injuries and deaths are reduced if active travel and public transport use are combined with urban structural renovation. The smart design approach champions a safe environment for walking and cycling.

Additional benefits:

- There is improved employee productivity leading to enormous financial gains with land use patterns that promote active travel.^16^ This is due to the physical and mental health benefits.
- Empowers vulnerable groups like women, children, the elderly, and those with low socioeconomic status who have poor access to private vehicles and supports them to lead independent lives.^16^
- The economic benefits from increased physical activity and reduced air pollution due to switching to active travel and public transport use were quantified by a study in Austria. It was estimated to be between 12-19 million € (approximately £10 to 16 million as of 20^th^ August 2022). If intangible costs such as those due to pain, anxiety, and hypothetical values of life lost are included, the most modest estimation is between 350-740 million € (£300 to 630 million). And the more optimistic estimates range between 910 million to 2.5 billion € (£770 million to 2.1 billion).^49^

There is significant evidence that the effects of climate change are not equally borne by different population groups. Such as the elderly (individuals aged 65 years and above) have been recognised as a highly climate-vulnerable group. The physiological abilities of the human body decline with age resulting in deterioration of respiratory, cardiovascular, and thermoregulatory functions. These functions are often worsened by pre-existing co morbidities such as diabetes, obesity, and hypertension. And within this subgroup, there is further variation in the effects borne due to the social determinants of health, such as low socioeconomic status, poor access to healthcare services, and inadequate housing infrastructures that limits their ability to adapt. Evidence also suggests that it is often difficult for the elderly to drive or own a car (due to diminished sensory awareness & alertness, living with specific disabilities and financial constraints) with a higher dependence on public transport. Thus, the public transport network in London needs to be strengthened to ensure all regions of the borough have accessibility to affordable public transportation services. Additionally, these transport services should be able to accommodate those living with a variety of disabilities (such as those with specialised medical equipment). Further, to encourage cycling, E-bikes must be promoted among the elderly as they might struggle with conventional cycling due to reduced stamina and other physical ailments. People (especially women, children, the elderly, and the disabled) who walk or cycle on roads are at higher risks of transport-related injuries and are considered vulnerable. This risk can be mitigated through strategic infrastructure planning that prioritises active transport over motorised transport. As evidence shows that improved transport and urban planning are the most effective means of encouraging physical activity.^16^

Active travel should be the main form of transport for most of the population, as it has almost zero GHG emissions, helps improve air quality, and has additional health co-benefits such as reduction of obesity-related diseases and improved mental health due to increased physical activity. It is also an equitable mode of transport, as costs associated with walking or cycling are minimum. Replanning neighbourhoods with healthy land usage can ensure active travel is the natural transportation choice. Disincentivising private car ownership through speed restrictions, a higher parking levy, and car-free neighbourhoods can encourage active travel for short-distance journeys and public transport use for long-distance trips. London has a well-connected and intricate public transport network which can be further strengthened by expanding to other regions currently underserved by public transport. Regular use of private electric vehicles should be discouraged, barring exceptions, such as those unable to actively travel, use public transportation, or deliver goods and other emergency services. This is because there are substantial particulate emissions from the tyres and brakes of EVs.^11,13^ Despite the commitments by the UK government to allocate 1.2 billion pounds to implement active travel, there have been budget cuts, with only 1/4^th^ of that amount available to the local authorities.^2^Ironically, there has been a simultaneous increase in investment to develop and expand motorised roads.^2^

Although measures such as the congestion charges have helped reduce GHG emissions and improve the air quality in London^61^, they are regressive as such inequitable strategies are eventually ineffective with higher income groups (who almost always have the highest carbon footprints). However, the revenues generated can improve health equity if invested in enhancing & subsidising public transport services and developing active travel-promoting environments. Providing bicycles in the most deprived areas through bike sharing schemes that do not require smartphones or credit cards can help improve health equity. For example, the Santander cycles currently offered by the TfL require a credit/debit card or a smartphone app to hire a bicycle.^68^ The process of hiring these bicycles could be made more seamless and equitable if these bicycles could be rented using the Oyster travel cards, which are currently only used to commute via bus, trains, or the tube network. Additionally, investments to provide access to affordable E-bikes can overcome many hindering factors associated with conventional cycling, such as difficulty riding up hilly terrains or poor physical fitness.^69^

### 5.4 Limitations of the study

One of the major limitations of assessing health risks addressed, and health equity impacts of transport strategies through the CAPs is that most boroughs might have mentioned them more elaborately in a separate document specific to that area. For example, exclusive air quality, heatwave, flooding, or food security action plans. Further, some documents like adaptation plans might already be in the pipeline, but the discussion and recommendations made in this report are shaped upon the finding that currently, there is minimal climate adaptation action.

This report has not assessed the implementation of the strategies outlined in the CAPs. There needs to be a formal evaluation, which was beyond the scope of this report. However, this could be a topic of consideration for further study. Additionally, many CAPs were written and published before the commencement of the COVID-19 pandemic. The aspirations and priorities of the councils might have substantially changed, leading to the evolution of climate-related actions. Such as, the importance of health and equity through climate actions might have marginally increased post the pandemic. But this needs to be evaluated as well and has not been done as a part of this report.

The recommendations made in this report are based on the evidence synthesised from analysis of the CAPs, interviews, and other scientific literature (which included recent systematic reviews). However, a formal literature review has not been conducted due to time constraints but could potentially make the recommendations more robust.

The sample of the qualitative element of the study is small, and the views and perspectives of the participants are not generalisable findings. However, the objective, which was to develop an in-depth understanding and provide insight into the role of health and equity in developing CAPs through qualitative interviews^18^, has been achieved. Many interviewees mentioned that there is a process for equality impact assessments within the local authorities. These might have been separately done and hence were not a part of the CAPs. So, this limits the conclusions drawn about health equity considerations in the CAPs. An equality impact assessment was found for the borough of Brent along with the CAP and provided better insight. But it makes the comparisons of health equity assessments in transport strategies outlined in the CAPs amongst the boroughs non-uniform. The researcher conducted, transcribed, coded, and analysed the interviews. This could have led to the potential single researcher bias in the findings of this report. To minimise this bias, the researcher revisited the qualitative data collected from the interviews after a certain period with a fresh perspective. There is also potential participant bias as the interviewees were aware of the research topic, which might have resulted in them shaping their answers around health equity.

> “But you need equity (in active travel). And obviously, your focus is also on equity, which I think is really interesting. And again, you said you want to produce some recommendations. That’s the normal thing in a health policy report.” (P3)

### 5.5 Implications for policy and practice

The following recommendations are being made to the GLA and the local authorities to ensure that health equity is an integral part of climate action by the local authorities. These recommendations are based on findings from the review of all the CAPs, current scientific evidence, and interviews of key officials from the London boroughs. A point to be noted is that these recommendations are mindful of the ambitious target of London becoming a net zero carbon city by 2030. Therefore, they would require a substantial financial investment and intense commitment to deliver results. Further, some of these recommendations could lead to initial public objection (such as active transport being the natural mode of transportation for the majority of the population) but needs to be innovatively and strategically tackled to turn the non-acceptance into approval.

#### 5.5.1 Recommendations to improve the role of health equity in climate action plans of the London boroughs

- Increasing public health capacity in climate action – It is essential to have a dedicated public health team member for climate action-related work. This requires additional financial investments and resources to upskill the public health task force.
- Conducting Joint strategic needs assessment (JSNA) for climate change – Systematic JSNAs should be a requisite for all London boroughs to generate evidence for effective climate action. This would be possible by increasing the role of public health specialists in climate emergency-related work.
- Lobby for incorporating climate change-related health indicators and outcomes into the Public Health Outcome Framework (PHOF) – The public health team frequently utilises this. Hence, it is crucial to have accessible climate change-related health data available at the fingertips. This could be under the realm of the broader health protection indicator of the PHOF.
- A public health climate change dashboard should be developed – It would contain all the climate-related information such as current borough-wise data, evidence, tools, and resources available in one place. This would aid local authorities in developing equitable strategies to improve health through climate action.
- Adaptation plans need to be formulated and implemented immediately – Utilisation of an evidence-based tool like the Local Climate Adaptation Tool (LCAT) can help incorporate strategies focusing on health and equity. The adaptation plans should be integrated and aligned with mitigation plans to minimise trade-offs.
- Strategic communication with different stakeholders – The opportune use of immediate and visible effects of climate change, such as during heatwaves, can garner public support for climate action. Stakeholders should be notified of the financial and health co-benefits in addition to environmental benefits that can be achieved through the implementation of mitigation and adaptation strategies.
- Evaluation of the CAPs - Process and impact evaluations of the CAPs are needed to ensure whether the objectives have been met and assess the implications of the strategies on population health and equity.

#### 5.5.2 Recommendations to maximise health and equity benefits from transport strategies aimed at reducing carbon emissions

- Incorporate ‘Win-Win’ strategies - To maximise the benefits to human health, equity and the planet ‘win-win’ actions such as making active travel the natural mode of transport for short distances and public transport for long distances. To foster an environment for active travel to thrive, smart redesigning of urban form such that all essential amenities are within walking or cycling distance and green spaces are abundant.
- Present evidence about the macroeconomics of the co-benefits of specific climate action to policymakers – To help argue in favour of health equity through climate action, information such as returns to investment in active travel should not be calculated in terms of just reduction of carbon emission, but also health gains through improved air quality, increased physical activity, and its positive impact on equity.

## 6. Conclusion

The local councils aim to achieve improved health and equity through climate action, but this needs to be better reflected in the current strategies. The current climate action plans of the different London boroughs have varied levels of integration of health into their strategies. But overall, health is sparsely integrated into climate action. And while some of the boroughs have considered equitable transport strategies like active travel with possible significant health and other co-benefits, there needs to be a renovation of infrastructural and environmental support.

Increasing public health capacity in climate action within the local councils and ensuring a ‘Health in all policies’ approach can help form and implement equitable strategies to combat climate change. Focusing on ‘Win-Win’ solutions in the transport sector would maximise long-term gains in terms of health, equity, environment, and finance.

## Data Availability

All data produced in the present study are available upon reasonable request to the authors

